# Exploring the Diagnostic Potential of miRNA Signatures in the Fabry Disease Serum: A Comparative Study of Automated and Manual Sample Isolations

**DOI:** 10.1101/2024.03.25.24304836

**Authors:** Josephine Y. Fang, Saravanan Ayyadurai, Alyssa F. Pybus, Hiroshi Sugimoto, Mark G. Qian

## Abstract

Fabry disease, an X-linked lysosomal storage disorder caused by galactosidase alpha (GLA) gene mutations, exhibits diverse clinical manifestations, and poses significant diagnostic challenges. Early diagnosis and treatment are crucial for improved patient outcomes, pressing the need for reliable biomarkers. In this study, we aimed to identify miRNA candidates as potential biomarkers for Fabry disease using the KingFisher™ automated isolation method and NanoString nCounter® miRNA detection assay.

Clinical serum samples were collected from both healthy subjects and Fabry disease patients. RNA extraction from the samples was performed using the KingFisher™ automated isolation method with the MagMAX mirVana™ kit or manually using the Qiagen miRNeasy kit. The subsequent NanoString nCounter® miRNA detection assay showed consistent performance and no correlation between RNA input concentration and raw count, ensuring reliable and reproducible results. Interestingly, the detection range and highly differential miRNA between the control and disease groups were found to be distinct depending on the isolation method employed. Nevertheless, enrichment analysis of miRNA-targeting genes consistently revealed significant associations with angiogenesis pathways in both isolation methods. Additionally, our investigation into the impact of enzyme replacement therapy on miRNA expression indicated that some differential miRNAs may be sensitive to treatment.

Our study provides valuable insights to identify miRNA biomarkers for Fabry disease. While different isolation methods yielded various detection ranges and highly differential miRNAs, the consistent association with angiogenesis pathways suggests their significance in disease progression. These findings lay the groundwork for further investigations and validation studies, ultimately leading to the development of non-invasive and reliable biomarkers to aid in early diagnosis and treatment monitoring for Fabry disease.

## Introduction

Fabry disease (FD, OMIM 301500) is a rare X-lined lysosomal storage disorder caused by the deficiency or malfunction of the enzyme α-galactosidase A(α-gal-A, EC3,2.1.22), resulting in the accumulation of globotriaosylceramide (Gb3) and related glycosphingolipids within lysosomes in various cells, including capillary endothelial, renal (podocytes, tubular cells, glomerular endothelial, mesangial and interstitial cells), cardiac (cardiomyocytes and fibroblasts) and nerve cells [1]. This progressive lysosomal storage disorder predominantly affects males and can lead to severe complications in the heart, kidneys, skin, eyes, central nervous system, and gastrointestinal system [2]. Due to the lyonization process inactivating one X-chromosome, some female patients have milder disease progression or delayed disease onset [3]. Despite advancements in FD research, the identification of reliable and specific biomarkers that reflect the disease progression and the response to treatment remains a challenge. Traditional methods, such as enzyme assays and genetic testing, have limitations in terms of sensitivity, specificity, and the ability to reflect clinical outcomes. Thus, there is a critical need for further research to identify and validate more useful biomarkers for FD.

Circulating miRNAs in liquid biopsies, such as serum samples, offer a minimally invasive, easily accessible source and are stable in long-term storage at room temperature, variable pH, and multiple freeze-thaw cycles, common conditions necessary for biomarker applications [4–7], making them an optimal option for identifying disease-specific molecular signatures for FD diagnostics. Current FD associated miRNAs have been reported about their key roles in cardiac function[8–10] and kidney disease progression[11–13]. However, the complex pathophysiology of FD and the low abundance of circulating miRNAs present additional hurdles for biomarker discovery. The NanoString nCounter® assay addresses these challenges by offering an image-based multiplex detection of miRNA sequences. Its digital counting capability ensures an accurate quantification, while the direct measurement of miRNAs without PCR amplification enhances sensitivity, minimizes variability, and avoids potential biases towards abundant miRNA from RNA amplification [14, 15]. Furthermore, this platform has received FDA 510(k) clearance and is currently in use for testing the PAM50 gene signature to assess the risk of recurrence in breast cancer and patient stratification in postmenopausal women [16, 17]. Therefore, a Nanostring-based miRNA assay platform may provide clinical utility.

Recently, Cammarata et al. have demonstrated the feasibility of using Nanostring assay with manual (Qiagen miRNeasy Serum/Plasma kit) miRNA isolation to identify common plasma miRNA profiles in FD patients[18]. However, the reported assay process is labor intensive and subject to variations from operators in conducting miRNA isolations. As shown in this work, we successfully interfaced the Nanostring assay with an automated process for isolation of the total small RNA by leveraging the capabilities of a KingFisher™ automation system (MagMAX™ total RNA isolation kit). The combination of the KingFisher™ automation system and NanoString nCounter® assay could accelerate the workflow to assess miRNA expression profile in serum samples. Using the improved assay workflow, we identified distinct miRNA signatures associated with the occurrence of disease and the response to enzyme replacement therapy (ERT) with a detailed comparison of miRNA profile between manual and automation isolations. This established automated assay and procedure are generic and could be readily applicable for creating reliable assays to monitor the complex excretion profile of miRNA in any liquid biopsies for FD diagnosis, prognosis, and evaluation of treatment outcomes.

## Methods and materials

### Patient and control of serum samples

Fabry patient serum samples were obtained from Sanguine Bioscience (Waltham, MA) following the Institutional Review Board (IRB) approved protocol (Project number 09286, No. San-BB-02). Healthy control serum samples were purchased from Discovery Life Sciences (Huntsville, AL), and individuals within a similar age range (±10 years) were selected to match the patient samples (Table 1). The samples were collected from December 5, 2017 to November 2, 2022 according to vendor’s documentation.

**Table 1.** Demographics of the study population.

| **Manual Method** |  |  |  |  |
| --- | --- | --- | --- | --- |
|  |  | **Total(n=24)** | **Healthy(n=10)** | **Fabry(n=14)** |
| **Age** | Median (years) | 52.5±13.55 | 51.0±14.23 | 53.0±13.55 |
|  | Ranges(years) | 21-66 | 21-65 | 29-66 |
| **Genders** | Male | 12 | 5 | 7 |
|  | Female | 12 | 5 | 7 |
| **Enzyme replacement treatment (ERT)** | No | 15 | 10 | 5 |
|  | Yes | 9 | 0 | 9 |
| **Automated Method** |  |  |  |  |
|  |  | **Total(n=23)** | **Healthy(n=12)** | **Fabry(n=11)** |
| **Age** | Median(years) | 50.0±14.26 | 56.5±14.43 | 47.0±14.68 |
|  | Ranges(years) | 21-75 | 21-70 | 31-75 |
| **Genders** | Male | 13 | 4 | 9 |
|  | Female | 10 | 8 | 2 |
| **Enzyme** | No | 12 | 12 | 2 |

| replacement |  |  |  |  |
| --- | --- | --- | --- | --- |
| treatment (ERT) | Yes | 7 | 0 | 7 |

RNA isolation and purification

Serum samples were thawed on ice and 200 µL of each sample was transferred into new tubes. miRNAs were isolated using miRNeasy Serum/Plasma Kit (cat. no. 217184, Qiagen, MD). Some samples were alternatively extracted using KingFisher™ Flex Magnetic Particle Processor 96DW with MagMAX mirVana™ Total RNA Isolation Kit (cat. no. 5400630 and A2728, both from ThermoFisher Scientific, MA), following the manufacturer’s protocol. For comparison purposes, eight samples were proceeded using both manual and machine-based methods. The miRNA concentrations were quantified using Qubit™ microRNA assay Kits (ThermoFisher Scientific, MA) with Qubit™ Flex Fluorometers.

### miRNA expression profiling and data analysis

The miRNA expressions were profiled using NanoString nCounter® with the Human v3 miRNA CodeSet kit (CSO-MMIR15-12) with nCounter® miRNA Sample Prep (Mu-MIRTAG-12). The codeset consists of 827 unique miRNAs barcodes, including six positive controls, eight negative controls, six ligation controls, five spike-in controls (ath-miR-159a, cel-miR-248, miR-254, and osa-miR-414 and 442). Additionally, five reference controls (housekeeping mRNAs: RPL10, ACTB, B2M, GAPDH, and RPL19) were included. Input material comprised 5 µL of concentrated RNA, and the experiment was run according to the manufacturer’s protocol. An illustrated experiment workflow is shown in Fig 1.

**Fig 1.**
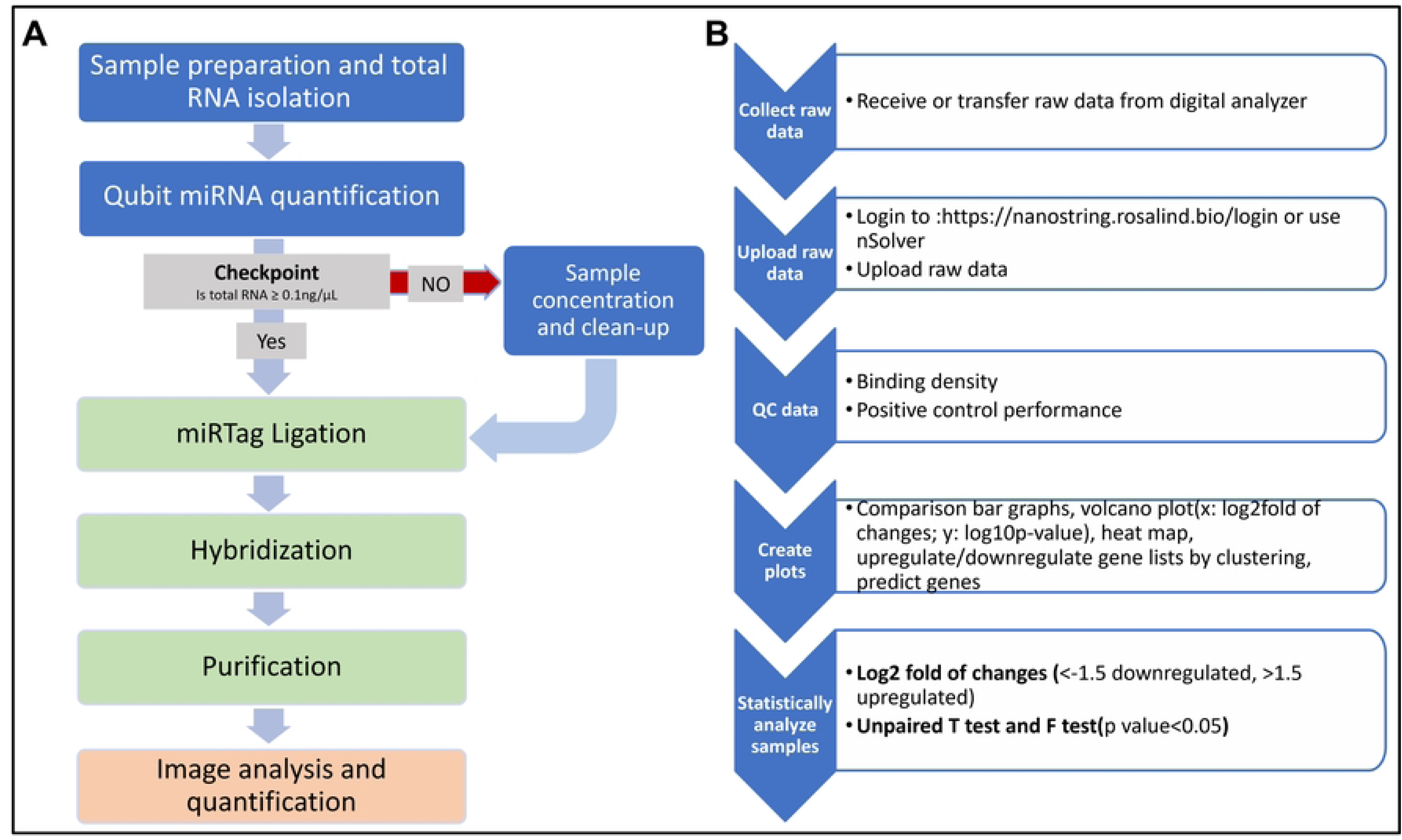
Workflow for Nanostring-based miRNA biomarker assay. (A)The flowchart illustrates the overall experimental procedures. The workflow consists of RNA sample preparation steps(blue), Nanostring nCounter sample preparation steps(green), and an image analysis step to obtain raw count data(orange). (B) The flowchart presents the data analysis steps after receiving the raw data.

All hybridizations were carried out for 19 hours, and data counts were obtained by scanning on the MAX model for 555 fields of view (FOV) per sample. The normalization of data (RCC file) was performed using nSolver Analysis Software v4.0 (NanoString Technologies, https://nanostring.com/products/analysis-solutions/ncounter-analysis-solutions/). Background correction was performed by subtracting the geometric mean of negative control counts by sample within NanoString nSolver™. Positive control and CodeSet normalization utilized the geometric mean of positive controls and the top 100 highly expressed miRNAs probe set according to nSolver™ guidelines. Fold change expression differences were calculated using nSolver Ratio data based on normalized count data. The data was further filtered for miRNA measured above detection levels in at least 15% of samples. Subsequent data analysis was performed using Rosalind (https://nanostring.rosalind.bio/login). Rosalind conducts differential expression analysis using the R package limma to obtain fold change values and p-values [19]. Heatmaps of differentially expressed miRNAs from Rosalind were scaled and generated using Ward’s hierarchical agglomerative clustering method with the fpc R package [20]. Volcano plots were created using R packages ggplot2, and ggrepel to customize figure sizes and ranges. The cut-off criteria for significance were set at |log2(fold of change)| > 1.5 and p-value < 0.05.

### Prediction and validation of miRNA-Target Interactions

To predict and validate the miRNA-target interactions of the highly differential miRNAs between Fabry and healthy control groups, we used MiRWalk (http://mirwalk.umm.uni-heidelberg.de). We employed the following default parameters: species human, miRNAID, GeneSymbol, Statistic power: 0.95, Position: 3UTR. The databases used for prediction were TargetScan, miRDB, and miRTarBase, and only interactions listed in all three (plus miRWalk’s own database) were included [21].

### Gene set enrichment analysis (GSEA) and functional annotation

Gene set enrichment analysis (GSEA) was performed to identify significantly enriched functions of the predicted target genes. System analysis, annotation, and visualization of gene function were conducted using the Reactome, the Kyoto Encyclopedia of Genes and Genomes (KEGG), and Gene ontology enrichment analysis. Pathways achieving an FDR (False Discovery Rate)-corrected p<0.05 were considered statistically significant. GSEA results were summarized in a bubble plot.

## Results

### RNA yield and NanoString counts from different isolation methods

To expedite and improve the sample processing workflow, we compared the manual and automated miRNA extraction methods by a correlation analysis by NanoString raw counts and miRNA input concentration. The sum of NanoString raw counts derived from the manual method ranged from 19,003-71,662 with a median of 29,637, whereas the automation method detected raw counts ranging from 15,860-37,857 with a median of 18,494. The Pearson correlation test R^2^ value for the overall dataset, manual method, and automation method were all lower than 0.3 (overall: 0.21; manual and automation: 0.056, Fig 2). These results suggest that miRNA input concentration did not significantly influence the total NanoString raw counts.

**Fig 2.**
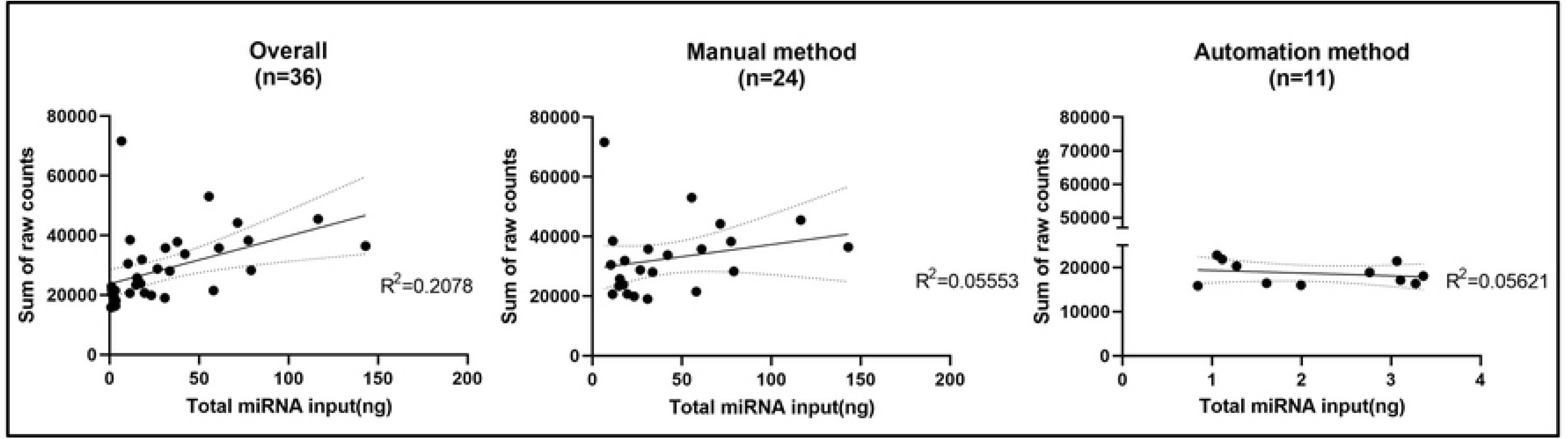
The correlation analysis of total miRNA input concentration and the sum of counts in samples from different isolation methods. Pearson correlation with linear regression analysis was performed. Mean linear regression is plotted (black straight line).

Furthermore, we analyzed the miRNA detection range from different isolation methods in the same set of samples in different cartridges (n=8). On average, 225±29 miRNAs were commonly detected in both methods across samples. Additionally, 123±69 miRNAs were exclusively detected in the manual method, while 58±40 miRNAs were exclusively detected in the automation method. The variations in individual microRNA species were depicted through average counts in both automated and manually isolated samples (Fig 3). The normalized counts of each miRNA from different methods were correlated to each other in a scatter plot, and the majority of dots aligned with the expected value (assuming no difference between manual and automation labeled as a green line in Fig 3). However, a subset of outliers displayed higher average counts in the manual method (orange dots closer to the x-axis). Seventy-three percent of the total detected miRNAs displayed higher normalized counts in the manual isolation method, while the remaining 27% presented higher counts in the automated approach. To identify significantly different microRNA species between isolation methods, we conducted a two-tailed Welch’s t-test, paired by the patient, comparing normalized counts of either method. Consequently, we identified 14 miRNAs that were significantly higher in the manual isolation method compared to the automated isolation method. These observations indicate that the profile of detected miRNAs is substantially influenced by the type of isolation method.

**Fig 3.**
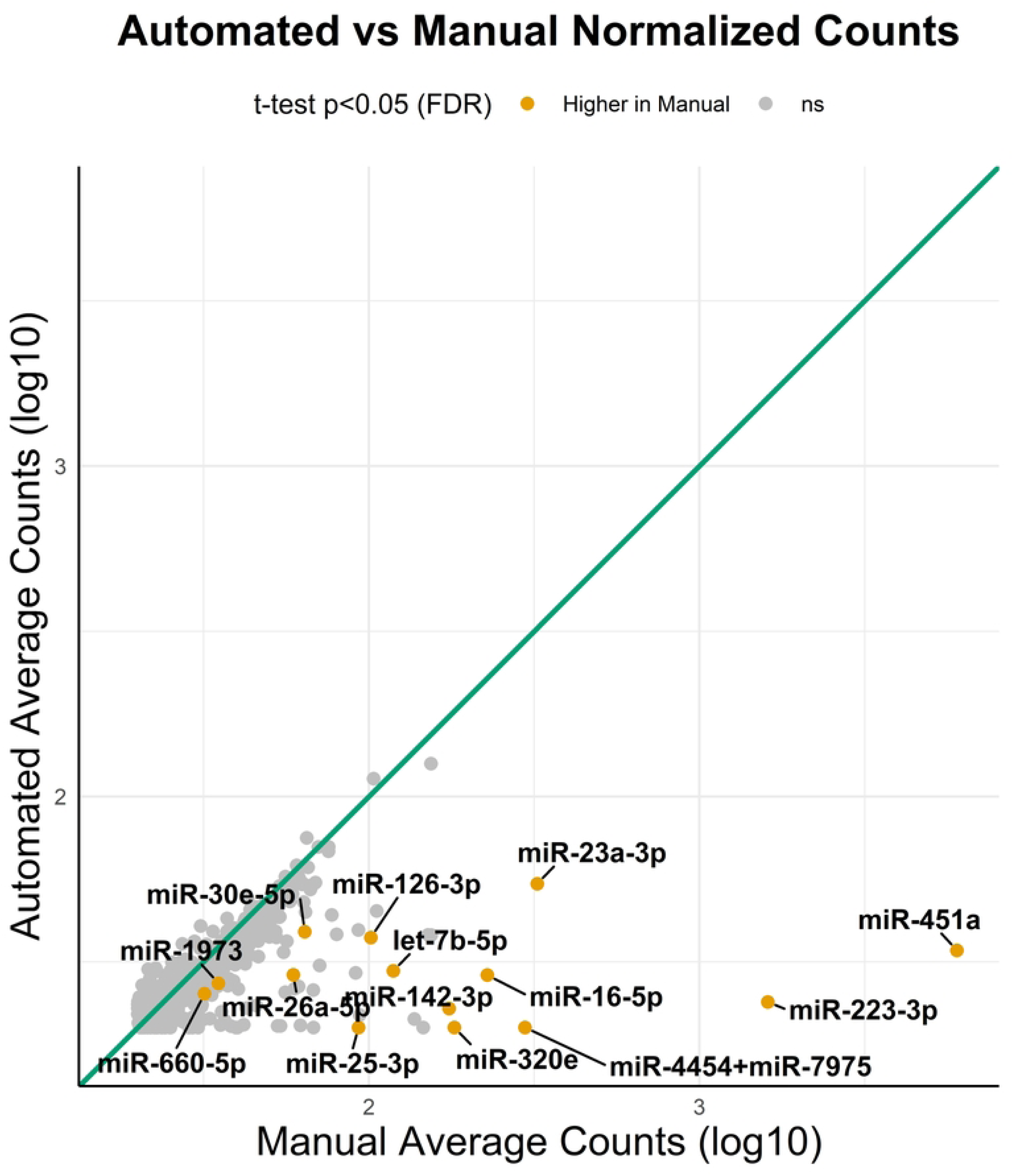
A scatter plot of normalized counts of both automated and manual isolation methods. The normalized counts from the 8 samples that have isolated miRNAs from both methods. A theoretical line (green line) indicating normalized counts of miRNAs were similar in both methods. Orange dots represent miRNAs displayed higher counts in the manual than in automated methods(p<0.05).

### Assay performance and quantification parameters

To assess method-derived variations, we compared miRNA profiles between healthy and FD groups using RNA samples extracted via manual and automated methods. A total of 39 serum samples were analyzed, consisting of 21 males and 18 females. Among these, only eight samples were processed using both methods. Table 1 provides a summary of the age range, gender distribution, and enzyme replacement treatment (ERT) status of the subjects.

The RNA concentration of manually extracted samples ranged from 1.33-28.6 ng/µL (median of 5.76 ng/ µL) while automated samples had lower concentrations ranging from 0.13-0.72 ng/µL (median of 0.47 ng/µL). To ensure assay quality, positive control linearity, binding density, and ligation efficiency were assessed (S1 Fig). Before evaluating these quantification parameters, we defined three levels of stringency for the background as the limit of detection (LOD): Low LOD=geometric mean of all negative controls; Medium LOD = geometric mean of negative control+2*standard deviation; High LOD = 2*(Medium LOD). The three levels of LOD in this assay were 21, 36, and 72 for the manual method and 21, 35, and 70 for the automated method, respectively.

In the manual method, control raw counts ranged from 91 to 31,663, while the automated method showed control raw counts ranging from 114 to 33,665 (S1A and 1D Figs). Correlation analysis of the raw counts and the positive control concentration (Pos_A-F) exhibited significant linearity with R^2^≥0.95 (S1B and S1E Figs, p<0.0001) in both methods. In contrast, binding density showed no correlation (R^2^≤0.3) with RNA input concentration (S1C and S1F Figs). The linear curve of the positive controls was used for assuring assay quality and the Low LOD was used as the criterion to assess the detection range of miRNAs.

Ligation efficacy was evaluated using three positive and three negative synthetic RNA controls. Ideally, the ligation-negative control should yield counts in the Low and Medium LOD range, while the ligation-positive should yield counts significantly higher than all three LOD levels. In the manual method, all ligation-negative controls were lower than Medium LOD, with an average of 4 out of 24 samples falling below the Low LOD (S1C Fig). In the automated method, negative controls were lower than Medium LOD in the majority of samples, except one, with an average of 6 out of 23 samples below the Low LOD (S1F Fig). All ligation-positive controls were higher than the Low LOD, except one sample in the manual method and eight samples in the automated method, which were lower for LIG_POS_C. When compared to the Medium LOD, the majority of samples were higher, with 1 or 3 out of 24 samples in the manual method and 7 or 9 samples in the automated method falling below the Medium LOD. A similar trend was observed for the High LOD, with an average of 4 out of 24 samples in the manual method and 8 out of 23 samples in the automated method below the High LOD. These quality control results provide essential information regarding assay performance and define criteria for normalization and quantification in data analysis.

### Differential miRNA detection ranges and expression profiles

To investigate the detection range of miRNA in different study groups isolating from either manual or automated methods, we employed a Low LOD threshold, with miRNAs considered detectable if present in at least 15% of samples. Across the serum sample, a total of 501 miRNAs were detected using the manual method, and 492 miRNAs were detected using the automated method. In the manual method, 100 miRNAs were exclusively detected in the Fabry group, 4 miRNAs were exclusively detected in the healthy group, and 397 were detected in both groups (Fig 4A). The automated method revealed 24 miRNAs solely detected in the Fabry group, 112 miRNAs exclusively detected in the healthy group, and 356 miRNAs detected in both groups (Fig 4B). Notably, 15 highly differential miRNAs were identified in the manual method, comprising 9 downregulated (green dots) and 6 upregulated (red dots) miRNAs that met the predefined criteria (Fig 4C). Among these 15 miRNAs, only 4 were specific to the Fabry group, while the remaining 11 miRNAs were commonly detected in both groups. In contrast, the automated method identified 29 highly differential miRNAs, consisting of 8 downregulated (green dots) and 21 upregulated (red dots) miRNAs (Fig 4D). Of these, only 1 miRNA was unique to the Fabry group, 3 miRNAs were unique to the healthy group, and the other 25 miRNAs were expressed ubiquitously in both groups.

**Fig 4.**
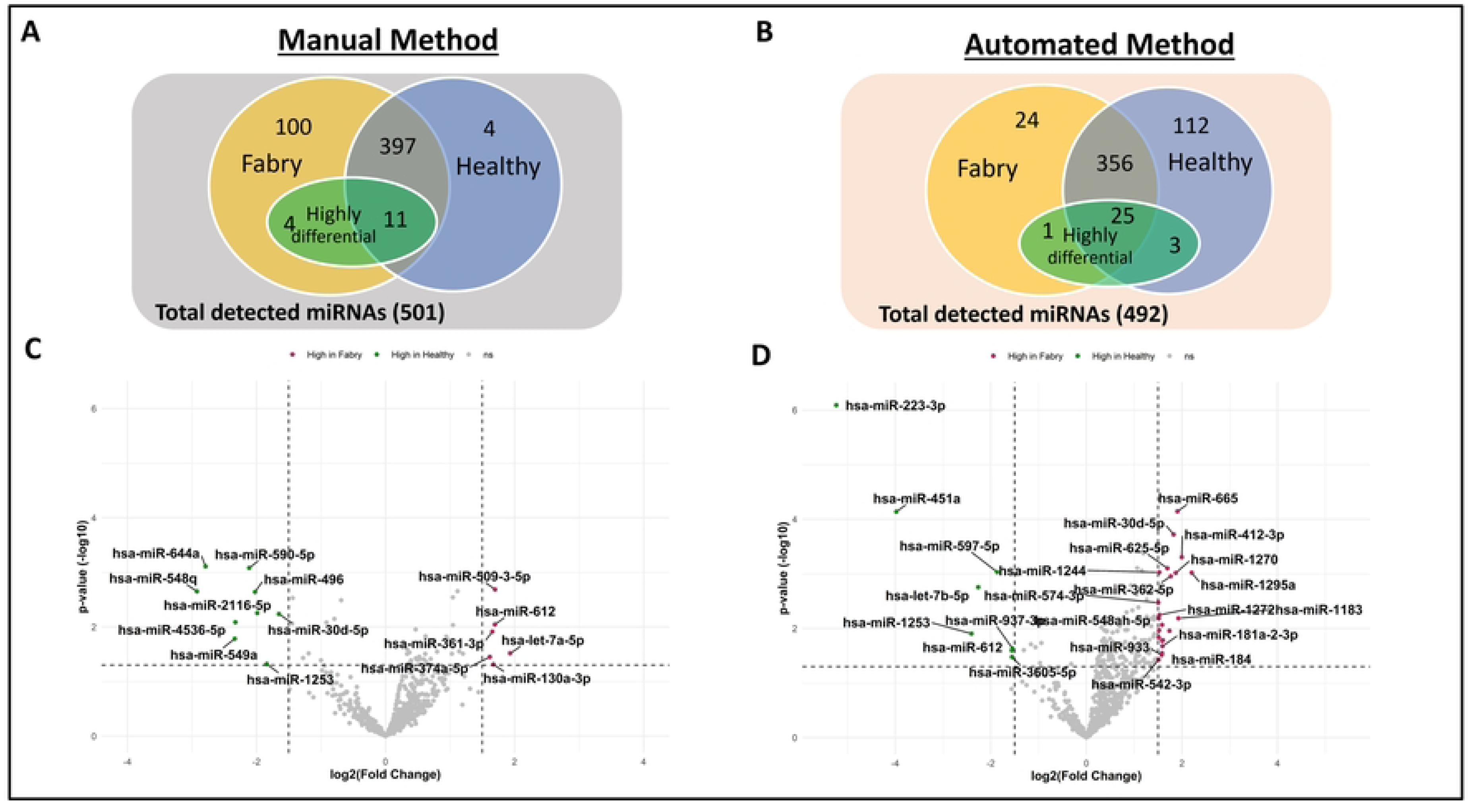
The miRNA detection range and highly differential miRNAs in healthy and Fabry serum samples from different isolation methods. (A and B) Venn diagram displays the detection range of the Fabry(yellow) and healthy(blue) groups. The green circle indicates the highly differential miRNAs within either Fabry or commonly expressed miRNAs regions. (C and D) Volcano plot shows log2 fold change(y-axis) and p-value(x-axis) across miRNAs. Colors indicate the differential level of the miRNA, either downregulating(green), or upregulating(red).

The overall distribution of differentially expressed miRNAs was visualized using a Volcano plot, with a significance threshold set at p-value<0.05 and |log2 foldchange|>1.5. In both the manual and automated methods, the sample dots were more densely populated in the positive region of the x-axis than the negative (Figs 4C and 4D). In the comparison between the manual and automated methods, we observed different miRNA expression patterns. Specifically, in the manual approach, six miRNA were found to be significantly elevated in Fabry samples compared to healthy samples (miR-509-3-5p, miR-612, miR-361-3p, let-7a-5p, miR-130a-3p, miR-374a-5p), whereas nine miRNA showed significant downregulation (miR-644a, miR-590-5p, miR-548q, miR-496, miR-2116-5p, miR-4536-5p, miR-30d-5p, miR-549a, miR-1253). In the automated method, 14 miRNA were elevated in the Fabry group compared to healthy controls (miR-665, miR-412-3p, miR-30d-5p, miR-1270, miR-1295a, miR-625-5p, miR-362-5p, miR-181a-2-3p, miR-184, miR-542-3p, miR-933, miR-548ah-5p, miR-1244, miR-574-3p), while 8 miRNA were reduced (miR-612, miR-597-5p, let-7b-5p, miR-1253, miR-451a, miR-223-3p, miR-937-3p, miR-3605-5p). Notably, miR-30d-5p, miR-612, and miR-1253 were consistently detected in both isolation methods. Both the Venn diagram and volcano plot results suggested that FD may considerably increase the number of detectable miRNA expressions.

In the heatmap and hierarchical clustering analyses, the Fabry group samples were separated into three clusters when compared to the healthy group. Specifically, the Fabry samples, F1 and F3, obtained via the manual method exhibited a miRNA expression pattern similar to the healthy group and were distant from the other Fabry samples (Fig 5A). The middle cluster (F4, F5, F2, and F13) displayed a mixed miRNA expression pattern between disease and healthy groups. The remaining samples generally demonstrated a distinct miRNA expression pattern associated with FD. A similar clustering pattern was observed in the sample obtained via the automated method. Mainly, F21 showed a miRNA expression pattern similar to the healthy group, while F22, F23, F24, and F25 exhibited an intermediate miRNA pattern, and the final group displayed a more pronounced Fabry-specific miRNA expression pattern (Fig 5B). After decoding the background information of the samples, we observed that all non-ERT patients were clustered in the first two groups, which exclusively included female patients above age 53. Additionally, samples in these two groups consisted of male patients who received ERT, with ages either below or above 53. The results imply that miRNA expression profiles might be influenced by age, gender, and response to ERT.

**Fig 5.**
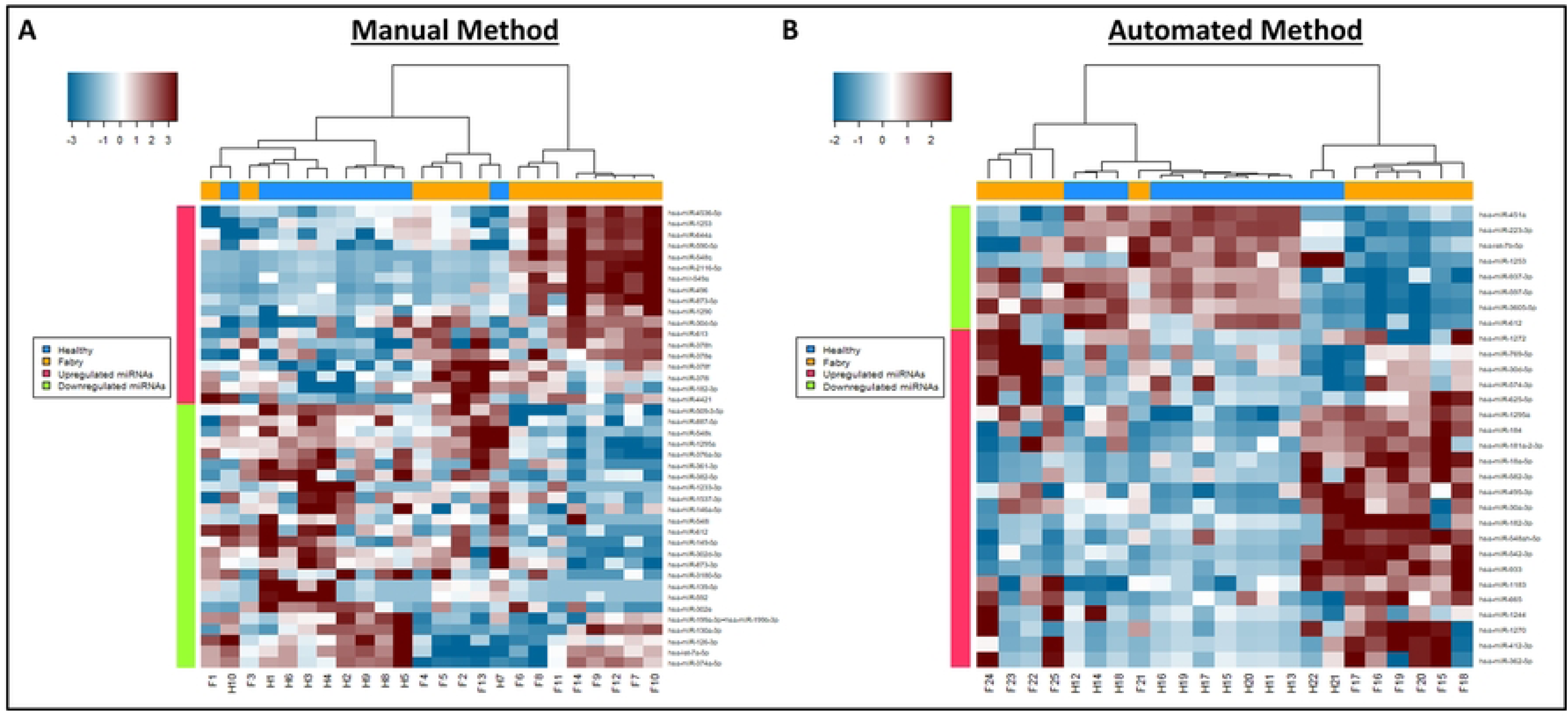
Hierarchical clustering heatmap for healthy and Fabry serum samples derived from different isolation methods. Hierarchical clustering heatmap of differential miRNAs (|log2 fold of change|>0.5 and p-value<0.05, total 42 miRNAs included) presents in each healthy control and Fabry patient serum samples. Colors encoded the up-(red) and down-(blue) regulated miRNAs.

### Prediction and annotation of a potential target of differentially expressed miRNAs

To identify potential target genes associated with highly differential miRNAs, we utilized miRWalk integrating TargetScan, miRDB, and miRTarBase to cross-reference miRNAs-target interactions. In the manual method, we identified a total of 211 miRNAs-target interaction sites involving miR-30d-5p, miR-130a-3p, miR-374a-5p, and let-7a-5p (Fig 6A). Similarly, in the automated method, we found 314 miRNAs-target interaction sites involving miR-18a-5p, let-7b-5p, miR-30d-5p, miR-665, miR-223-3p, miR-362-5p, and miR-495-3p (Fig 6B).

**Fig 6.**
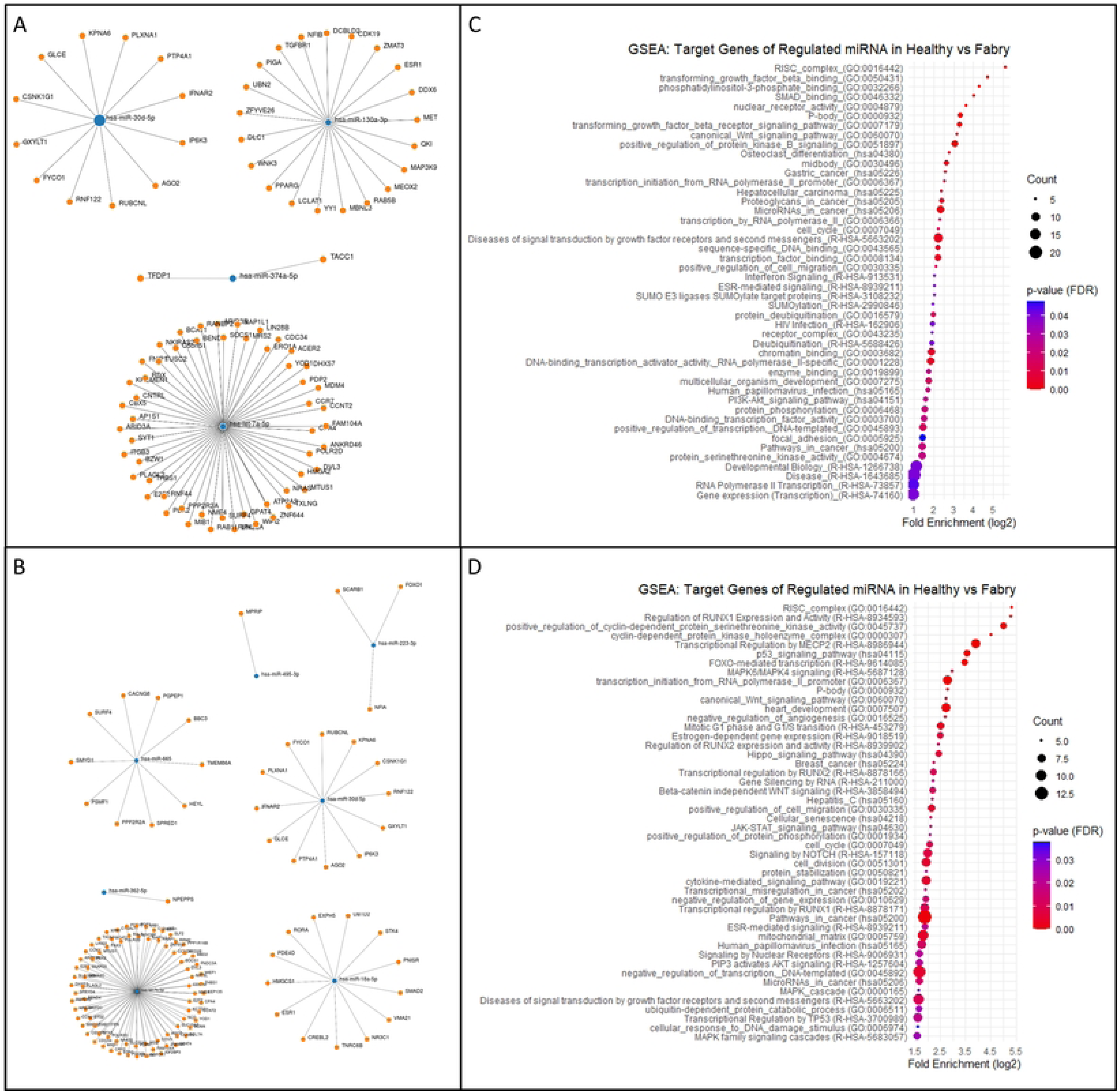
Predicted target genes and function enrichment analysis. (A and B) Node graphs display the miRNA (blue dot)-target (orange dot) interactions. Filters for target gene prediction were seed sequences mapping to the 3’ untranslated regions (UTRs), a p-value of 0.05, and only targets identified by three different algorithm-TargetScan, miRDB, and miRTarBase. (C and D) The bubble dot plot represents the fold enrichment (x-axis), the number of genes (bubble size), and the FDR value (gradient colors) in each GO and KEGG biological process terms (y-axis). Representative bubbles were enriched at FDR<0.05.

To gain insight into the biological function of the identified miRNA-associated target gene in Fabry serum samples, we conducted gene set enrichment analysis in miRWalk, including GO (Biological Process, Molecular Function, and Cell Component) and KEGG. In the manual-method-derived Fabry samples (Fig 6C), gene ontology analysis revealed significant enrichment in pathways such as TGF-β binding and receptor signaling, SMAD binding, PI3K-Akt signaling, and phosphatidylinositol-3-phosphate binding, which are known to be associated with FD [22, 23]. Similarly, gene set enrichment analysis on the automated-method-derived Fabry samples(Fig 6D) showed enrichment in pathways related to RUNX1 and RUNX2 regulations and activities, heart development, negative regulation of angiogenesis, and MAPK signaling pathways, which have been reported to be associated with various angiogenesis-related disorders such as cancer, cardiomyopathy, nephropathy, and retinopathy[24–27]. In addition to angiogenesis-related signals, the automated-method-derived Fabry samples displayed enrichment in the Notch signaling pathway, which is thought to be involved in kidney fibrosis in the FD [28]. These findings highlight the possibility that upregulated miR-30d-5p and downregulated let-7a-5p or let-7d-5p are associated with angiogenesis-related signaling pathways in various aspects.

### miRNAs expression profiles with and without enzyme replacement therapy (ERT)

To further evaluate the miRNA expression patterns between ERT-treated and non-ERT-treated Fabry patients, we focused on highly differentially expressed miRNAs identified from the comparison between healthy and non-ERT-treated Fabry patients and compared their expression level with the ERT-treated group. In the manual method, we identified a total of 29 highly differential miRNAs (|log2 fold change|>0.5 and p-value<0.05). Among these, 21 miRNAs displayed expression levels similar to those of the healthy group, indicating a potential recovery of their expression in response to ERT. Conversely, eight miRNAs showed no significant change compared to the non-ERT treated group (representative miRNAs, Fig 7A). Similar trends were observed with the automated method (representative miRNAs, Fig 7B). Employing the previously established highly differential criteria (|log2 fold change|>1.5 and p-value<0.05), we identified 21 highly differential miRNAs. Out of these, 17 miRNAs presented a tendency to approach the expression levels observed in the healthy group upon ERT treatment, whereas 4 miRNAs showed no significant change.

**Fig 7.**
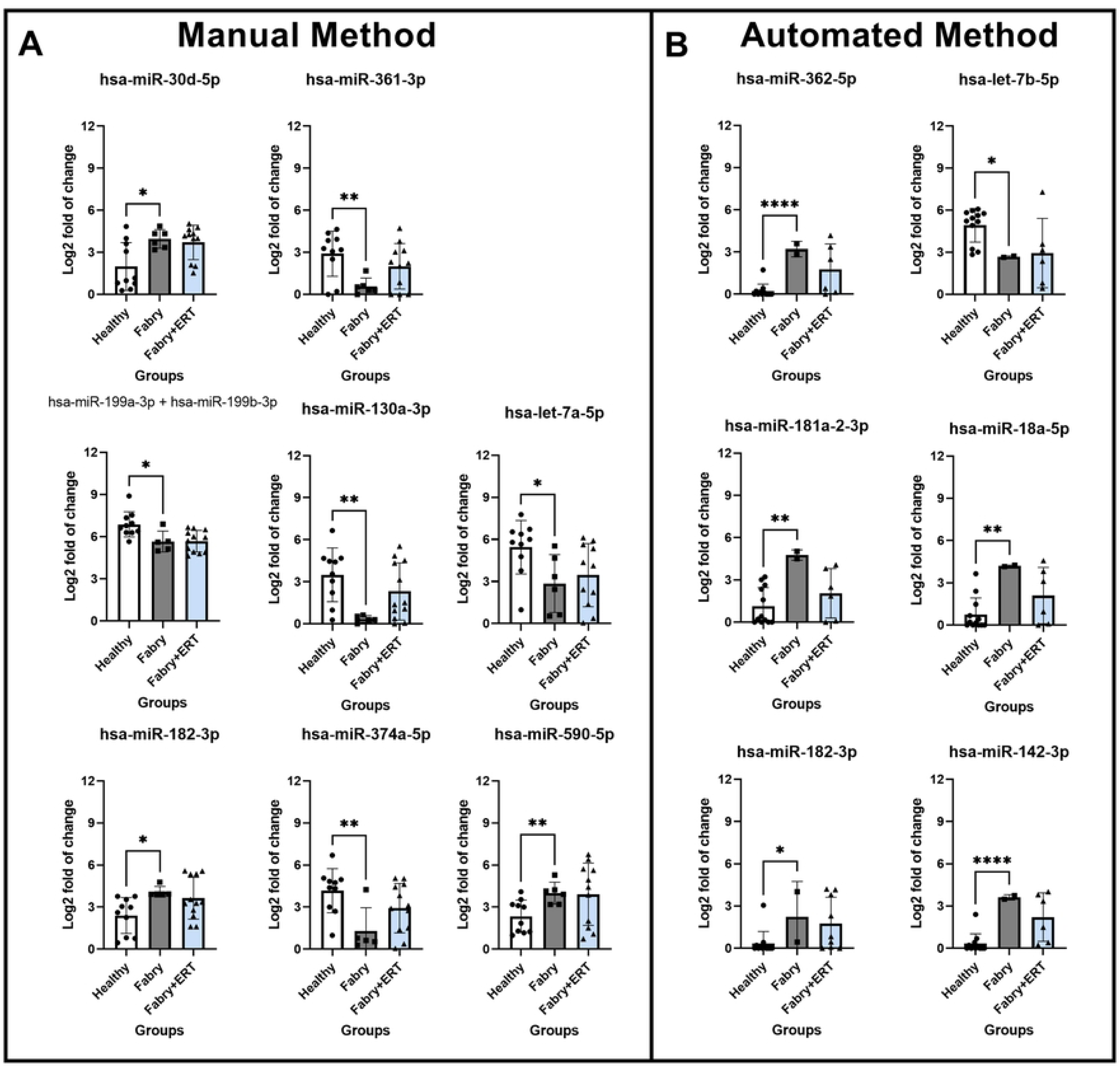
ERT impacts on the differentially expressed miRNAs associated with the Fabry pathology. Representative miRNAs were significantly differential between Healthy and non-treated Fabry group. Statistical comparison of mean performed by nonparametric T-test with p value<0.5*, 0.01**, 0.001***, 0.0001****.

Upon reviewing the relevant literature, we found that some of these differentially expressed miRNAs have known involvement in processes related to vascular formation, autophagy, and muscle differentiation [29–36]. The ERT improved the expression of most of these differential miRNAs toward the levels observed in the healthy group. The result suggests that these miRNAs could potentially serve as candidates to monitor disease progression and the effectiveness of therapeutics.

## Discussions

In this study, we investigated the impact of different isolation methods on miRNA expression profiles in Fabry patients using the NanoString nCounter® platform. We found that the isolation method did not significantly affect the assay performance and sum of NanoString raw counts. However, it did have an impact on the type of miRNAs detected and the list of highly differential miRNAs in the comparison between Fabry patients and healthy controls. Despite these differences, both methods consistently demonstrated that serum from Fabry patients presented greater miRNA diversity and more clustered miRNA expression profiles according to gender, age, and ERT status compared to the healthy control group, even with the divergence in the list of highly differential miRNA. These highly differential miRNAs are likely involved in a variety of pathways related to vascular formation, autophagy machinery, muscle homeostasis, and differentiation pathways [8, 9, 37–41].

The enrichment analysis provided quantitative evidence that the hub of genes derived from the highly differential miRNAs, regardless of the isolation method used, regulates different aspects of angiogenesis signals. Notably, previous research has linked the malfunction of TGF-β, PI3K, and Notch to Fabry’s disease nephropathy [1, 42, 43]. Moreover, we found that the majority of these differential miRNA expression levels could be altered by the ERT status. Therefore, these highly differential miRNAs could serve as a potential biomarker panel to track disease progression and treatment response as long as the results are obtained from the same isolation method.

A similar Fabry biomarker discovery study conducted by Giuseppe Cammarata et al. employed the same Qiagen miRNA isolation kit and NanoString nCounter® platform (v2 instead of v3 in our study)[18]. They identified 18 highly differential miRNAs (p-value<0.05, |log2 fold of change|≥1.5) when compared to healthy controls. In the comparison of our results (the manual method part), two miRNAs, miR-126-3p and miR-146a-5p, were commonly identified in both studies, but the value of log2 fold of change was smaller and regulating directions (up and down) were opposite in our results. Additionally, both studies identified miR-199 and miR-361, let-7, but the subtype (−3p or −5p and a or b) and the regulating directions were different. These inconsistencies might result from the demographics of the sample age and gender. Fabry’s disease is an X-linked genetic disorder, and female patients often reveal milder symptoms than male patients. Consequently, the clinical manifestation of disease limits the availability of samples, and in our study, all non-ERT treated patient serum samples were obtained from female patients above the age of 53. Age is known to contribute to the heterogeneity of the serum miRNA profile in Fabry patients, and it also impacts miRNA expressions in healthy individuals. For instance, we observed a strong correlation between miR-181a-5p and miR-499b-5p with age (R^2^=0.4988, p=0.0224 and R^2^=0.588, p=0.0097, S2 Fig). Thus, the differential miRNAs identified may vary depending on the sample resource, but ultimately, these miRNAs are likely to target pathways associated with vascular differentiation or formation.

In addition to the biomarker discovery, we also demonstrated possible parameters that could impact assay performance in the NanoString nCounter® assay when implementing an automated RNA isolation method. The variation in miRNA profiles was mainly due to the divergence of the isolation method rather than the RNA input concentration (Fig. 2). Similar results were observed in several studies on miRNAs in serum. For instance, Marjorie Monleau et al. compared miRNA profiles using different RNA isolation kits and found that the extraction procedure impacted the detection range and G/C composition of miRNAs [44]. Similarly, Ryan Wong et al. evaluated the detection range of miRNAs from two different RNA isolation kits (MagnaZol RNA and Qiagen kits) for RNA-sequencing and found differences in the reads mapping to miRNAs and the diversity of detected miRNAs [45]. Moreover, the choice of RNA fraction used in the miRNA microarray and bead array-based assays can also impact miRNA profiles [46, 47]. While there may be discrepancies in the miRNA detection range and the list of highly differential miRNA between automated and manual isolation methods, it is encouraging to note that a subset of miRNAs was consistently identified from both methods. This overlap adds robustness to the finding and strengthens the validity of the identified miRNAs as potential biomarkers for FD. Furthermore, the highly differential miRNA lists obtained through both methods demonstrated enrichment in angiogenesis-related pathways, which aligns with the biological context of the disease manifestation. The enrichment analysis provides important insights into the potential functional roles of these miRNAs in the pathogenesis of FD, particularly in relation to vascular formation and angiogenesis.

The current study has some limitations that need to be addressed. First, the small sample size and imbalanced gender and age distribution could introduce bias. However, the rarity and X chromosome-dependent nature of FD poses significant challenges in recruiting sufficient sample sizes with equally distributed genders and ages. Therefore, the selection of healthy control samples is crucial to match the disease group and reduce the impact of age on miRNA expression profiles during comparisons. Second, no single RNA kit can perfectly preserve all types of miRNAs for the experiment, resulting in a bias of miRNA detection range and the variation of target genes and pathways. Experiments performed in different cartridges and dates add another layer of variation within the data. Data comparisons should be performed optimally within the same experiment process to minimize the isolation method and batch bias. Although different isolation methods may yield different lists of highly differential miRNAs, the sample hierarchical clustering and functional analysis can still identify disease-associated pathways. Finally, there is no one-fit-for-all target prediction platform. Currently, most computational algorithms use classic seed pairing principles, which target-to-seed sequences within the 3’ untranslated regions (UTRs) of mRNA. However, miRNA can bind to regions beyond the 3’ UTRs, and seed base-paring does not necessarily need to be perfect, as it can be complemented by additional 3’ compensatory base-paring [23, 48–52]. To overcome the risk of over-prediction, we cross-validated computationally predicted miRNA-target interactions with miRTarBase, a database of experimentally validated interactions [53].

This study highlights the potential of automating the miRNA isolation process with the NanoString nCounter® assay for FD biomarker discovery. The implementation of this automated workflow offers several advantages over traditional manual methods. Firstly, it effectively reduces technical variations arising from hands-on experiment procedures, thereby enhancing the reliability and reproducibility of miRNA expression data. Moreover, the automation of the process reduces labor time, making it feasible to process a large number of samples simultaneously.

One potential method modification that can be considered is the choice of magnetic beads used in the RNA isolation protocol. Currently, magnetic beads-based RNA isolation protocol is specialized for total RNA [54]. However, it may be beneficial to develop magnetic beads that specifically target small RNAs to further enhance the isolation efficiency and accuracy. Such modification can potentially improve the miRNA detection range and better concordance between isolation methods.

In conclusion, our results demonstrated that an automated workflow for miRNA isolation with NanoString nCounter® assay could identify a panel of miRNAs targeting similar hubs of angiogenesis genes as the manual method. These miRNAs can potentially serve as biomarkers for diagnosis, prognosis, surveillance, and even in therapeutic applications. The workflow is applicable to investigate miRNA expression signatures associated with other diseases for biomarker studies.

## Data Availability

All data produced in the present study are available upon reasonable request to the authors and all data produced in the present work are contained in the manuscript. However, the data are available in Takeda internal electronic notebook system network.

## Acknowledgments

We acknowledged Lavesh Gwalani and Joshua Chi for their generous provision of clinical samples for our study and Yusuke Sato for generously sharing techniques and usages of Nanostring nCounter machines.

## Disclosure

JYF, SA, HS, and MGQ are current employees at Takeda and have ownership of Takeda stock. AFP was under employment of Takeda during the project. This work was funded by Takeda Development Center Americas Inc.

## Supporting information

**S1 Fig. Assay performance and quantification parameter.** A and D) Correlation analysis between raw counts (Log2) and positive control concentration (Log2. fm). A Simple linear regression of R2 was calculated for the data. B and E) Correlation analysis between binding density and RNA input concentration(ng). Pearson correlation with linear regression analysis was performed. Mean linear regression is plotted (black straight line) with 95% confidence intervals (dashed line). C and F) Digital counts (Log2) are shown for all ligation controls. Average Low LOD, Medium LOD, and High LOD thresholds calculated for all negative controls are highlighted in shades of blue.

**S2 Fig. Age impacts the miRNAs expression profile in healthy subjects.** The association between age and miRNAs expression was performed by Person Correlation Coefficient. Both P values were lower than 0.05.

## Notes

### Funding Statement

This study did not receive a specific funding.

### Author Declarations

IRB of Advarra and WCG gave ethical approvals for this work.

## References

1. Germain DP. Fabry disease. Orphanet journal of rare diseases. 2010;5(1):1–49.

2. Bernardes TP, Foresto RD, Kirsztajn GM. Fabry disease: genetics, pathology, and treatment. Revista da Associação Médica Brasileira. 2020;66:s10–s6.

3. Beck M. Demographics of FOS–the Fabry outcome survey. Fabry disease: perspectives from 5 years of FOS. 2006.

4. Mitchell PS, Parkin RK, Kroh EM, Fritz BR, Wyman SK, Pogosova-Agadjanyan EL, et al. Circulating microRNAs as stable blood-based markers for cancer detection. Proceedings of the National Academy of Sciences. 2008;105(30):10513–8.

5. Mráz M, Malinova K, Mayer J, Pospisilova S. MicroRNA isolation and stability in stored RNA samples. Biochemical and biophysical research communications. 2009;390(1):1–4.

6. McDonald JS, Milosevic D, Reddi HV, Grebe SK, Algeciras-Schimnich A. Analysis of circulating microRNA: preanalytical and analytical challenges. Clinical chemistry. 2011;57(6):833–40.

7. Li Y, Jiang Z, Xu L, Yao H, Guo J, Ding X. Stability analysis of liver cancer-related microRNAs. Acta Biochim Biophys Sin. 2011;43(1):69–78.

8. Chistiakov DA, Orekhov AN, Bobryshev YV. Cardiac-specific miRNA in cardiogenesis, heart function, and cardiac pathology (with focus on myocardial infarction). Journal of molecular and cellular cardiology. 2016;94:107–21.

9. Braun T, Gautel M. Transcriptional mechanisms regulating skeletal muscle differentiation, growth and homeostasis. Nature reviews Molecular cell biology. 2011;12(6):349–61.

10. Zacharewicz E, Kalanon M, Murphy RM, Russell AP, Lamon S. MicroRNA-99b-5p downregulates protein synthesis in human primary myotubes. American Journal of Physiology-Cell Physiology. 2020;319(2):C432–C40.

11. Levstek T, Vujkovac B, Cokan Vujkovac A, Trebušak Podkrajšek K. Urinary-derived extracellular vesicles reveal a distinct microRNA signature associated with the development and progression of Fabry nephropathy. Frontiers in Medicine. 2023;10:1143905.

12. Jaurretche S, Perez G, Antongiovanni N, Perretta F, Venera G. Variables associated with a urinary MicroRNAs excretion profile indicative of renal fibrosis in Fabry disease patients. International Journal of Chronic Diseases. 2019;2019.

13. Levstek T, Mlinšek T, Holcar M, Goričar K, Lenassi M, Dolžan V, et al. Urinary extracellular vesicles and their miRNA cargo in patients with Fabry nephropathy. Genes. 2021;12(7):1057.

14. Veldman-Jones MH, Brant R, Rooney C, Geh C, Emery H, Harbron CG, et al. Evaluating Robustness and Sensitivity of the NanoString Technologies nCounter Platform to Enable Multiplexed Gene Expression Analysis of Clinical SamplesEvaluation of NanoString Technologies nCounter Platform. Cancer research. 2015;75(13):2587–93.

15. Tsang H-F, Xue VW, Koh S-P, Chiu Y-M, Ng LP-W, Wong S-CC. NanoString, a novel digital color-coded barcode technology: current and future applications in molecular diagnostics. Expert review of molecular diagnostics. 2017;17(1):95–103.

16. Nielsen T, Wallden B, Schaper C, Ferree S, Liu S, Gao D, et al. Analytical validation of the PAM50-based Prosigna Breast Cancer Prognostic Gene Signature Assay and nCounter Analysis System using formalin-fixed paraffin-embedded breast tumor specimens. BMC cancer. 2014;14:1–14.

17. Administration F. PROSIGNA BREAST CANCER PROGNOSTIC GENE SIGNATURE ASSAY FDA 510(k) Premarket Notification: FDA Administration; 07/01/2014. Available from: https://www.accessdata.fda.gov/scripts/cdrh/cfdocs/cfpmn/pmn.cfm?ID=K141771.

18. Cammarata G, Scalia S, Colomba P, Zizzo C, Pisani A, Riccio E, et al. A pilot study of circulating microRNAs as potential biomarkers of Fabry disease. Oncotarget. 2018;9(44):27333.

19. Ritchie ME, Phipson B, Wu D, Hu Y, Law CW, Shi W, Smyth GK. limma powers differential expression analyses for RNA-sequencing and microarray studies. Nucleic acids research. 2015;43(7):e47-e.

20. Murtagh F, Legendre P. Ward’s hierarchical agglomerative clustering method: which algorithms implement Ward’s criterion? Journal of classification. 2014;31:274–95.

21. Sticht C, De La Torre C, Parveen A, Gretz N. miRWalk: an online resource for prediction of microRNA binding sites. PloS one. 2018;13(10):e0206239.

22. Rozenfeld PA, de los Angeles Bolla M, Quieto P, Pisani A, Feriozzi S, Neuman P, Bondar C. Pathogenesis of Fabry nephropathy: the pathways leading to fibrosis. Molecular genetics and metabolism. 2020;129(2):132–41.

23. Park S, Kim JA, Joo KY, Choi S, Choi E-N, Shin J-A, et al. Globotriaosylceramide leads to KCa3. 1 channel dysfunction: A new insight into endothelial dysfunction in Fabry disease. Cardiovascular research. 2011;89(2):290–9.

24. Lam JD, Oh DJ, Wong LL, Amarnani D, Park-Windhol C, Sanchez AV, et al. Identification of RUNX1 as a mediator of aberrant retinal angiogenesis. Diabetes. 2017;66(7):1950–6.

25. Bao M, Chen Y, Liu J-T, Bao H, Wang W-B, Qi Y-X, Lv F. Extracellular matrix stiffness controls VEGF165 secretion and neuroblastoma angiogenesis via the YAP/RUNX2/SRSF1 axis. Angiogenesis. 2022;25(1):71–86.

26. Safa A, Abak A, Shoorei H, Taheri M, Ghafouri-Fard S. MicroRNAs as regulators of ERK/MAPK pathway: a comprehensive review. Biomedicine & Pharmacotherapy. 2020;132:110853.

27. Lee SH, Manandhar S, Lee YM. Roles of RUNX in hypoxia-induced responses and angiogenesis. RUNX Proteins in Development and Cancer. 2017:449–69.

28. Sanchez-Niño MD, Carpio D, Sanz AB, Ruiz-Ortega M, Mezzano S, Ortiz A. Lyso-Gb3 activates Notch1 in human podocytes. Human molecular genetics. 2015;24(20):5720–32.

29. Jia K, Shi P, Han X, Chen T, Tang H, Wang J. Diagnostic value of miR-30d-5p and miR-125b-5p in acute myocardial infarction. Molecular medicine reports. 2016;14(1):184–94.

30. Li X, Du N, Zhang Q, Li J, Chen X, Liu X, et al. MicroRNA-30d regulates cardiomyocyte pyroptosis by directly targeting foxo3a in diabetic cardiomyopathy. Cell death & disease. 2014;5(10):e1479-e.

31. Cheng X, Liu Z, Zhang H, Lian Y. Inhibition of LOXL1-AS1 alleviates oxidative low-density lipoprotein induced angiogenesis via downregulation of miR-590-5p mediated KLF6/VEGF signaling pathway. Cell Cycle. 2021;20(17):1663–80.

32. Zhang J, Zhou Y, Huang T, Wu F, Pan Y, Dong Y, et al. FGF18, a prominent player in FGF signaling, promotes gastric tumorigenesis through autocrine manner and is negatively regulated by miR-590-5p. Oncogene. 2019;38(1):33–46.

33. Luo Z, Sun Y, Qi B, Lin J, Chen Y, Xu Y, Chen J. Human bone marrow mesenchymal stem cell-derived extracellular vesicles inhibit shoulder stiffness via let-7a/Tgfbr1 axis. Bioactive Materials. 2022;17:344–59.

34. Yu S, Tang X, Zheng T, Li S, Ren H, Wu H, et al. Plasma-derived extracellular vesicles transfer microRNA-130a-3p to alleviate myocardial ischemia/reperfusion injury by targeting ATG16L1. Cell and Tissue Research. 2022;389(1):99–114.

35. Huang Z-Q, Xu W, Wu J-L, Lu X, Chen X-M. MicroRNA-374a protects against myocardial ischemia-reperfusion injury in mice by targeting the MAPK6 pathway. Life sciences. 2019;232:116619.

36. Farina FM, Hall IF, Serio S, Zani S, Climent M, Salvarani N, et al. miR-128-3p is a novel regulator of vascular smooth muscle cell phenotypic switch and vascular diseases. Circulation research. 2020;126(12):e120–e35.

37. Fernández-Hernando C, Suárez Y. MicroRNAs in endothelial cell homeostasis and vascular disease. Current opinion in hematology. 2018;25(3):227.

38. Zhai H, Fesler A, Ju J. MicroRNA: a third dimension in autophagy. Cell cycle. 2013;12(2):246–50.

39. Guo H, Pu M, Tai Y, Chen Y, Lu H, Qiao J, et al. Nuclear miR-30b-5p suppresses TFEB-mediated lysosomal biogenesis and autophagy. Cell Death & Differentiation. 2021;28(1):320–36.

40. Corà D, Bussolino F, Doronzo G. TFEB signalling-related MicroRNAs and autophagy. Biomolecules. 2021;11(7):985.

41. Delbridge LM, Mellor KM, Taylor DJ, Gottlieb RA. Myocardial stress and autophagy: mechanisms and potential therapies. Nature Reviews Cardiology. 2017;14(7):412–25.

42. Jeon YJ, Jung N, Jung SC. Globotriaosylceramide and globotriaosylsphingosine mediated epithelial-to-mesenchymal transition in kidney cells: implication for Fabry disease nephropathy (600.1). The FASEB Journal. 2014;28:600.1.

43. Feriozzi S, Rozenfeld P. Pathology and pathogenic pathways in Fabry nephropathy. Clinical and Experimental Nephrology. 2021;25:925–34.

44. Monleau M, Bonnel S, Gostan T, Blanchard D, Courgnaud V, Lecellier C-H. Comparison of different extraction techniques to profile microRNAs from human sera and peripheral blood mononuclear cells. BMC genomics. 2014;15(1):1–11.

45. Wong RK, MacMahon M, Woodside JV, Simpson DA. A comparison of RNA extraction and sequencing protocols for detection of small RNAs in plasma. BMC genomics. 2019;20:1–12.

46. Podolska A, Kaczkowski B, Litman T, Fredholm M, Cirera S. How the RNA isolation method can affect microRNA microarray results. Acta Biochimica Polonica. 2011;58(4).

47. Gaarz A, Debey-Pascher S, Classen S, Eggle D, Gathof B, Chen J, et al. Bead array–based microRNA expression profiling of peripheral blood and the impact of different RNA isolation approaches. The Journal of Molecular Diagnostics. 2010;12(3):335–44.

48. Moretti F, Thermann R, Hentze MW. Mechanism of translational regulation by miR-2 from sites in the 5′ untranslated region or the open reading frame. Rna. 2010;16(12):2493–502.

49. Forman JJ, Coller HA. The code within the code: microRNAs target coding regions. Cell cycle. 2010;9(8):1533–41.

50. Wei J, Gao W, Zhu C-J, Liu Y-Q, Mei Z, Cheng T, Shu Y-Q. Identification of plasma microRNA-21 as a biomarker for early detection and chemosensitivity of non–small cell lung cancer. Chinese journal of cancer. 2011;30(6):407.

51. Didiano D, Hobert O. Perfect seed pairing is not a generally reliable predictor for miRNA-target interactions. Nature structural & molecular biology. 2006;13(9):849–51.

52. Chi SW, Hannon GJ, Darnell RB. An alternative mode of microRNA target recognition. Nature structural & molecular biology. 2012;19(3):321–7.

53. Huang H-Y, Lin Y-C-D, Cui S, Huang Y, Tang Y, Xu J, et al. miRTarBase update 2022: an informative resource for experimentally validated miRNA–target interactions. Nucleic acids research. 2022;50(D1):D222–D30.

54. Rio DC, Ares M, Hannon GJ, Nilsen TW. Guidelines for the use of RNA purification kits. Cold Spring Harbor Protocols. 2010;2010(7):pdb.ip79.

